# Risk factors and outcomes of emerging Listeria monocytogenes infection in Pakistan: insights from a tertiary care hospital

**DOI:** 10.1101/2025.09.16.25335854

**Authors:** Zara Nadeem, Iffat Khanum, Imran Ahmed, Tahir Munir, Kiren Habib

**Author notes:** **Corresponding Author** Iffat Khanum: FCPS (IM) FCPS (ID), FRCP, MHPE, Assistant Professor, Dept. of Medicine, Aga Khan University, Stadium Road, Karachi 74800, Pakistan,00923333346194, Fax No: NA.

## Abstract

**Background:** *Listeria monocytogenes* is a Gram-positive rod responsible for listeriosis. This systemic infection often presents with bacteremia and may progress to severe manifestations such as meningoencephalitis, particularly in immunocompromised individuals and the elderly. In pregnant women, it is associated with fetal and placental involvement, frequently resulting in adverse outcomes. Over the past few decades, *L. monocytogenes* has emerged as a significant foodborne pathogen, implicated in numerous outbreaks worldwide. The objective of this study is to assess the risk factors, clinical features, and outcomes of patients admitted with *Listeria monocytogenes* infection at a tertiary care Centre in Karachi, Pakistan.

**Methods:** A retrospective study was conducted over 7 years and included all patients with culture-proven listeriosis. Comorbid conditions, clinical presentation, treatment, and outcomes were recorded and analyzed.

**Results:** A total of 63 patients diagnosed with *Listeria monocytogenes* infection were included in the study. There was female predominance (n = 44, 70.9%), with a mean age of 50.2 years. Diabetes mellitus was the most common comorbidity (43.5%). Among high-risk groups, 14 (22.6%) patients were pregnant, 10 (15.9%) were on immunosuppressive therapy, and another 10 (15.9%) were classified as elderly. The predominant presenting symptoms included fever (74.6%) and central nervous system involvement (52.0%), mainly meningoencephalitis. All patients received antibiotic therapy with either ampicillin or meropenem for a mean duration of 16.7 ± 8.4 days. The overall mortality rate was 11.1%.

**Conclusion:** Listeriosis was observed not only in the elderly but also in middle-aged individuals with underlying risk factors, as well as in pregnant women. Enhanced environmental hygiene, early diagnosis, and timely treatment are essential to improving outcomes, particularly in pregnancy-related cases. Public education, healthcare provider training, and community-level preventive strategies are critical for effective management and control of listeriosis.

## INTRODUCTION

*Listeria monocytogenes* is a gram-positive rod, known to be an opportunistic food-borne pathogen worldwide [1].Its most common route of transmission in the community is consumption of contaminated food[2]. *Listeria* causes food-borne illness, can survive even in low-moisture, low temperatures usually found in refrigerators, high salt foods making it challenging to control in food environment[3].Due to its prolonged incubation period, prompt diagnosis is difficult. Numerous reports of listeriosis outbreaks have been linked to dairy products, meat products, as well as fresh produce [4]. Apart from causing non-invasive, non-fatal infections, it was discovered as a major human pathogen causing fatal illnesses, including severe meningoencephalitis and maternal-fetal infections [5].

The annual incidence rate of listeriosis varies from 0.1 to 10 cases per million people per year, depending on the countries and regions of the world [6]. Ready-to-eat dairy, seafood, and immunocompromised individuals, including pregnant women and geriatric populations, seem to be most affected by this pathogen [7-9]. Despite the low prevalence, it has a high mortality rate, particularly in high-risk populations such as neonates, pregnant females, age ≥ 60 years, primary bacteremia, CNS involvement and prior comorbidities such as malignancies, chronic kidney diseases[10]. Maternal infections with *Listeria monocytogenes* have been reported to be rare before 20 weeks of gestations. The incidence of listeriosis is higher among pregnant women than in the general population and is associated with adverse fetal outcomes, including miscarriage and stillbirth[9, 11].

Treatment recommendations for listeriosis rely on knowledge gained from animal models, in vitro experiments, and clinical studies, as listeriosis is not a common disease and there aren’t many effective randomized controlled trials. Ampicillin is the recommended treatment, and trimethoprim/sulphamethoxazole,meropenem can be an alternative agent in case of penicillin allergy [12].Although penicillin and ampicillin are the recommended treatments for listeriosis, most experts advise combining gentamicin with ampicillin due to its delayed bactericidal activity against Listeria[13]. While some retrospective studies have failed to show a survival benefit— and even suggested potential harm in neurolisteriosis—other data indicate a possible survival advantage when gentamicin is administered for more than three days[14].

We aimed to identify the risk factors, clinical characteristics, and outcomes of listeriosis at a tertiary care centre, Pakistan. with the goal of understanding various clinical manifestations and predisposing conditions in our population to facilitate early recognition and management of this potentially treatable condition

## METHODOLOGY

This retrospective, observational study was conducted from January 2017 to December 2023 at the Aga Khan University Hospital (AKUH), Karachi, Pakistan. AKUH is a Joint Commission International (JCI) accredited state-of-the-art tertiary care centre that provides care to patients from all over the country. All patients admitted with culture-proven *L. monocytogenes* were evaluated for possible inclusion in the study. Patients with co-infections with other bacterial infections were excluded. An extensive review of electronic and paper-based medical records, together with radiology and laboratory databases of the selected patients, was undertaken. Pertinent information—including demographics, comorbid conditions, predisposing factors, immune status, imaging findings, management strategies, complications, and clinical outcomes—was documented on a structured proforma.

### Ethical consideration

The study was reviewed and approved by the Ethics Review Committee, Aga Khan University (ERC # 2023-9294-26813). Due to the retrospective chart reviews and lack of direct involvement of patients or other human participants, a waiver of informed consent was given by the Ethics Review Committee, Aga Khan University.

Data analysis was performed using RStudio (version 4.1.2; Boston, USA). The Shapiro-Wilk test was used to assess the normality of quantitative variables, including age, premature birth, and gestational age. As all variables were found to be non-normally distributed, they were summarized using median and interquartile range (IQR). Categorical variables, such as gender, comorbidities, clinical manifestations, and patient outcomes, were summarized as frequencies and percentages. Stratified analysis was conducted based on the diagnosis Neurolisteriosis vs. Non-neurolisteriossis to explore potential statistical associations. The Mann-Whitney U test was used for continuous variables, while the Chi-square or Fisher’s exact test was applied for categorical variables, as appropriate.

## RESULTS

A total of 63 patients met the inclusion criteria and were included in the study. The mean age of participants was 50.2 (Median 53, [Q1, Q3] [32.0, 67.0]) years, and the majority were female (n=44, 69.8%). Diabetes Mellitus (n=27, 43.5%) was the most frequent co-morbid illness, followed by hypertension (n= 25, 40.3%) and CLD (n=12, 19.0%) (Table 1). Among the high-risk group,14 (22.6%) patients were pregnant, and 10 (15.9%) patients were on immunosuppressive therapy, and a similar percentage was classified as elderly.

**Table 1:** Listeriosis and Comorbid Conditions.

| **Hypertension** | N | Percentage |
| --- | --- | --- |
| No | 38 | (59.7%) |
| Yes | 25 | (40.3%) |
| **Diabetes** |  |  |
| No | 36 | (56.5%) |
| Yes | 27 | (43.5%) |
| **HIV-Status** |  |  |
| No | 62 | (98.4%) |
| Yes | 1 | (1.6%) |
| **Autoimmune disease** |  |  |
| No | 56 | (88.9%) |
| Yes | 7 | (11.1%) |
| **Malignancy** |  |  |
| No | 57 | (90.5%) |
| Yes | 6 | (9.5%) |
| **Chronic Kidney Disease** |  |  |
| No | 58 | (92.1%) |
| Yes | 6 | (7.9%) |
| **Chronic Liver Disease** |  |  |
| No | 51 | (81.0%) |
| Yes | 12 | (19.0%) |
| **Others/ Description of known disease** |  |  |
| No | 33 | (52.4%) |
| Yes | 30 | (47.6%) |
| **Immuno-compromised** |  |  |
| No | 46 | (73.0%) |
| Yes | 17 | (27.0%) |
| **On Immunosuppressive therapy** |  |  |
| No | 53 | (84.1%) |
| Yes | 10 | (15.9%) |
| **Pregnancy Status** |  |  |
| No | 30 | (77.8%) |
| Yes | 14 | (22.2%) |
| **Extreme of Age** |  |  |
| No | 53 | (84.1%) |
| Yes | 10 | (15.9%) |
| **Other** |  |  |
| Alcoholic | 2 | (3.2%) |
| Hypothyroidism, hemolytic anemia | 1 | (1.6%) |
| Multiple myeloma | 1 | (1.6%) |

**Table 2:**
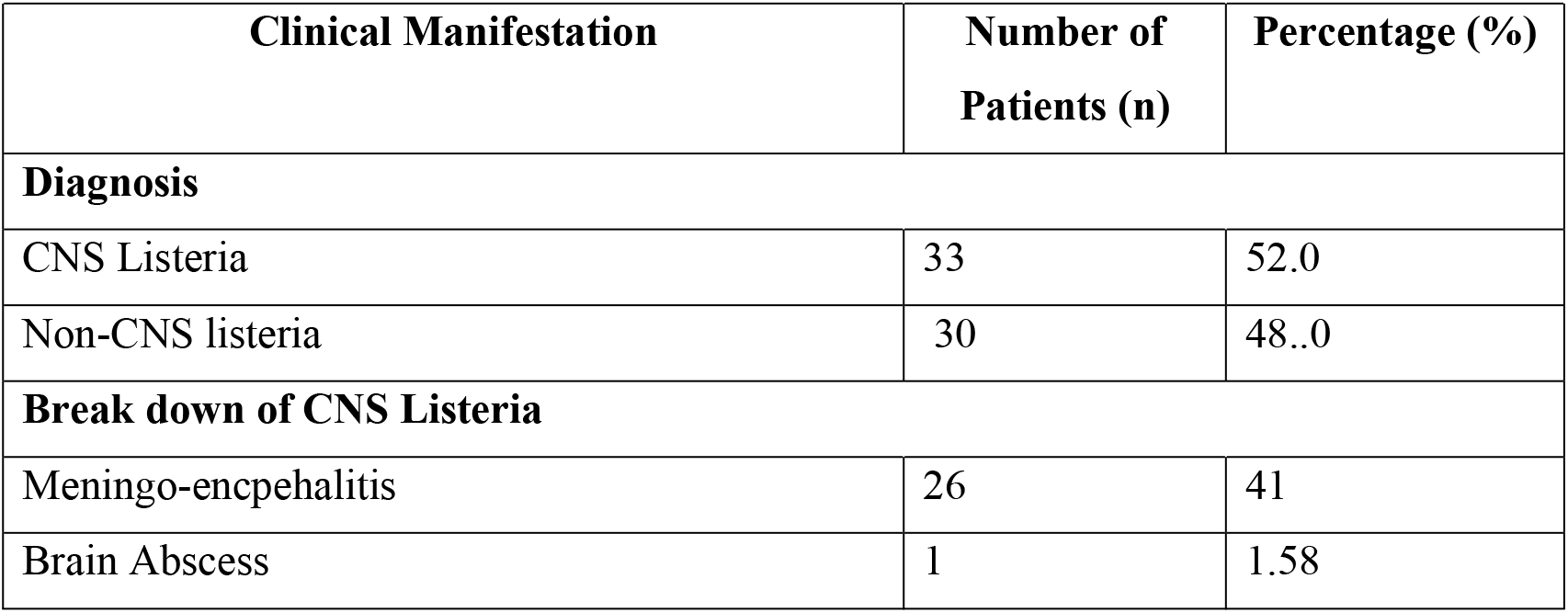
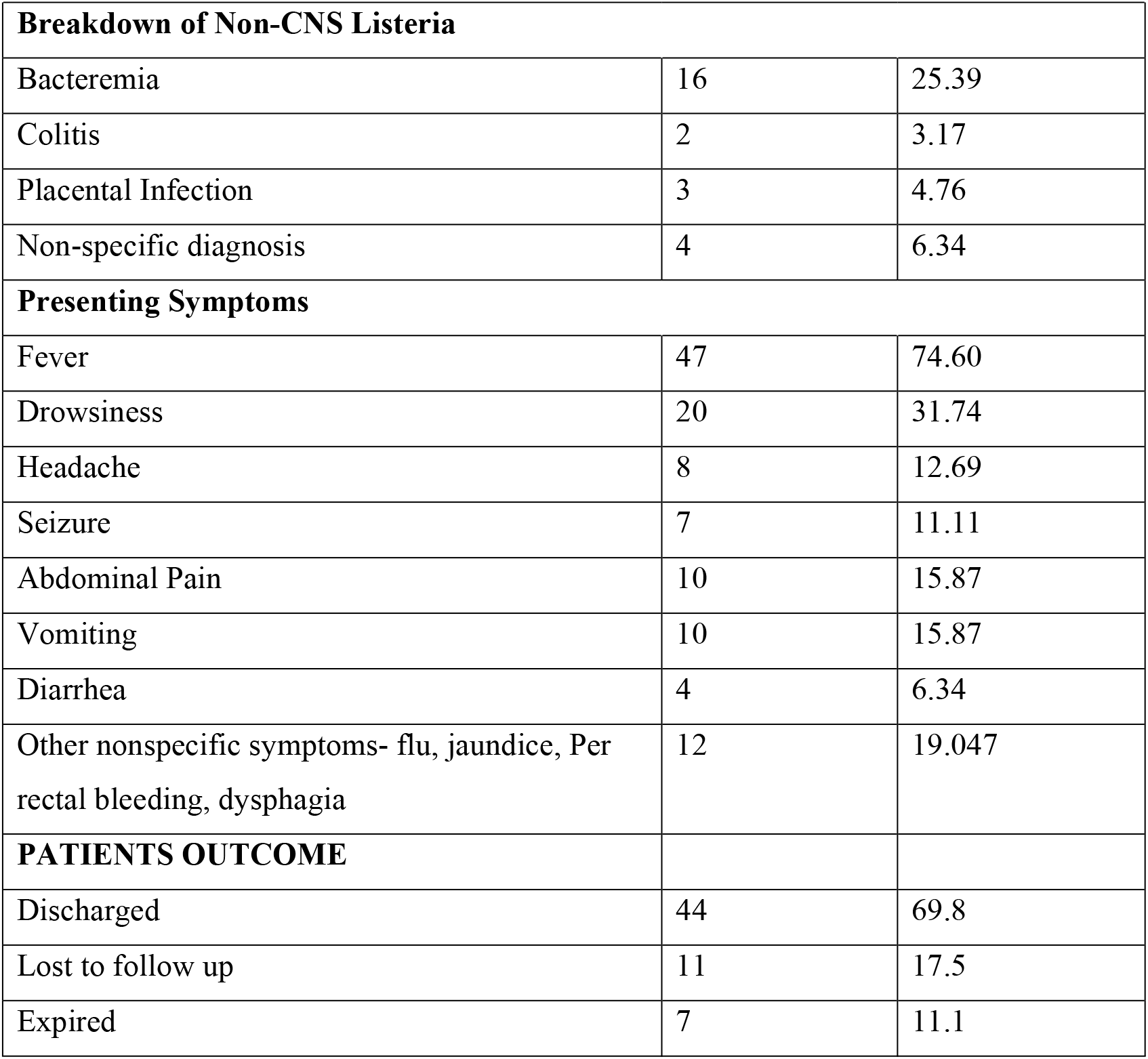
Clinical Manifestation of patients with Listeria Monocytogenes infection.

The most common clinical features were fever (n=47, 74.6%), followed by drowsiness (n=20, 31.7%), abdominal pain and vomiting (n=10, 15.9%). Nearly half (n=27, 52.0%) of the study patients presented with CNS listeriosis, predominantly meningoencephalitis, one patient had brain abscess, and 16 (25.39%) patients had concomitant bacteremia as well (Table 3). Overall, antimicrobial susceptibility data were available for 59 cases. In three cases where *Listeria monocytogenes was* detected by a commercial multiplex PCR (Filmarray meningitis encephalitis panel, Biomerieux) the organism did not grow on culture. The susceptibility rates for ampicillin, meropenem and trimethoprim/sulphamethoxazole were 96.6%, 100% and 86.4% respectively.

**Table 3:** Stratification analysis based on diagnosis.

| **Variable** | **CNS Listeria** | **Non-CNS listeria** | **P-value** |
| --- | --- | --- | --- |
|  | **(N=33)** | **(N=30)** |  |
| **Age (Years)** |  |  | 0.110 |
| Median [Q1, Q3Max] | 61.0 [35.0, 68.0] | 48.5 [29.5, 61.5] |  |
| **Gender** |  |  |  |
| F | 23 (69.7%) | 21 (70.0%) | 1.00 |
| M | 10 (30.3%) | 9 (30.0%) |  |
| **Hypertension** |  |  |  |
| No | 17 (51.5%) | 21 (70.0%) | 0.198 |
| Yes | 16 (48.5%) | 9 (30.0%) |  |
| **Diabetes** |  |  |  |
| No | 20 (60.6%) | 16 (53.3%) | 0.616 |
| Yes | 13 (39.4%) | 14 (46.7%) |  |
| **HIV Status- Affected** |  |  |  |
| No | 32 (97.0%) | 30 (100%) | 1.00 |
| Yes | 1 (3.0%) | 0 (0%) |  |
| **Autoimmune Disease** |  |  |  |
| No | 30 (90.9%) | 26 (86.7%) | 0.700 |
| Yes | 3 (9.1%) | 4 (13.3%) |  |
| **Malignancy** |  |  |  |
| No | 30 (90.9%) | 27 (90.0%) | 1.00 |
| Yes | 3 (9.1%) | 3 (10.0%) |  |
| **Chronic Kidney Disease** |  |  |  |
| No | 31 (93.9%) | 27 (90.0%) | 0.662 |
| Yes | 2 (6.1%) | 3 (10.0%) |  |
| **CLD** |  |  |  |
| No | 25 (75.8%) | 26 (86.7%) | 0.344 |
| Yes | 8 (24.2%) | 4 (13.3%) |  |
| **Others/ Description of known disease** |  |  |  |
| No | 17 (51.5%) | 16 (53.3%) | 1.00 |
| Yes | 16 (48.5%) | 14 (46.7%) |  |
| **Immuno-compromised** |  |  |  |
| No | 25 (75.8%) | 21 (70.0%) | 0.777 |
| Yes | 8 (24.2%) | 9 (30.0%) |  |
| **On Immunosuppressive** |  |  |  |
| No | 28 (84.8%) | 25 (83.3%) | 1.00 |
| Yes | 5 (15.2%) | 5 (16.7%) |  |
| **Pregnancy** |  |  |  |
| No | 13 (92.9%) | 7 (35.0%) | 0.001 |
| Yes | 1 (7.1%) | 13 (65.0%) |  |
| **Extreme of Age** |  |  |  |
| No | 26 (78.8%) | 27 (90.0%) | 0.308 |
| Yes | 7 (21.2%) | 3 (10.0%) |  |
| **Food Source** |  |  |  |
| No | 26 (78.8%) | 22 (73.3%) | 0.877 |
| NOT MENTIONED | 6 (18.2%) | 7 (23.3%) |  |
| Yes | 1 (3.0%) | 1 (3.3%) |  |
| **No Food Source** |  |  |  |
| No | 33 (100%) | 29 (96.7%) | 0.476 |
| Yes | 0 (0%) | 1 (3.3%) |  |
| **Socioeconomic status** |  |  |  |
| High | 11 (33.3%) | 11 (36.7%) | 0.639 |
| Low | 2 (6.1%) | 0 (0%) |  |
| Middle | 20 (60.6%) | 18 (60.0%) |  |
| Missing | 0 (0%) | 1 (3.3%) |  |
| **Other- Comorbid Conditions** |  |  |  |
| Alcoholic | 1 (3.0%) | 1 (3.3%) | 0.255 |
| Multiple myeloma | 1 (3.0%) | 0 (0%) |  |
| NA | 3 (9.1%) | 0 (0%) |  |
| No | 28 (84.8%) | 28 (93.3%) |  |
| Hypothyroidism, hemolytic anaemia | 0 (0%) | 1 (3.3%) |  |
| **If abortion/ premature birth, week of pregnancy** |  |  |  |
| Median [Q1, Q3] | 30.0 [25.0, 33.5] | 27.0 [24.0, 37.0] | 0.970 |
| **PATIENTS OUTCOME** |  |  |  |
| Discharged | 22 (66.7%) | 22 (73.3%) | 0.610 |
| Expired | 5 (15.2%) | 2 (6.7%) |  |
| Lost to follow up | 6 (18.2%) | 5 (16.7%) |  |
| **PREGNANCY OUTCOME** |  |  |  |
| EXPIRED | 2 (6.1%) | 5 (16.7%) | 0.024 |
| Premature delivery | 1 (3.0%) | 6 (20.0%) |  |

All patients were treated with ampicillin or meropenem, depending upon the availability of drugs for 4 – 6 weeks (mean 16.7±8.4) days. The overall mortality was 11.1% and most of the patients who died were older than 60 years of age. In subgroup analysis between CNS and non-CNS listeriosis (table 4), there was no major difference found, i.e age, comorbid illness and gender distribution and mortality. Most pregnant patients had non-CNS listeriosis (p value 0.014). Mortality was observed in 5 patients (15.2%) in CNS listeriosis and 2 patients (6.7%) in the non-CNS group. Pregnancy-related mortality was significantly higher in non-CNS involvement, with 5 patients (16.7%) compared to 2 patients (6.1%) in CNS listeriosis (p = 0.024).

## DISCUSSION

Our study showed that although listeriosis is common at extremes of age, it also significantly impacts the middle-aged population, especially those with diabetes and also pregnant females. Central nervous system (CNS) listeriosis was found to be as frequent as non-CNS cases. Although elderly patients were not the most frequently affected group, they accounted for the majority of the deaths, with an overall mortality rate of 11%.

Listeriosis is more common among patients with prior risk factors like pregnancy, immunosuppressive state or advanced age. A higher incidence of *L. monocytogenes* infection has been reported among individuals over 65 years of age, likely attributed to a weakened immune response and reduced ability to combat infectious agents.[15, 16]. The median age of our study population was 53 years, with only 15.9% patients being more than 65 years of age. Yan Liu et al. also found a median age of 56 years, but with 40.7% patients aged ≥60 years.[17] . In developing countries like Pakistan, increased cases of listeriosis among patients under 65 years of age may be secondary to poor food safety, inadequate sanitation, limited public awareness, weak health infrastructure and challenges in disease surveillance, diagnosis and prevention.[18, 19]

Pakistan faces a growing burden of diabetes mellitus (DM), increasingly affecting younger individuals. [20, 21] DM impairs immune responses, predisposing patients to infections. In our study, most patients had DM, which likely explains the lower median age (<65 years) of listeriosis cases compared to global patterns involving older adults.

Pregnant women are at increased risk of developing listeriosis than the general population, increased risk of severe disease, complications and adverse obstetric outcomes [22]. This is consistent with our study, in which 50% of pregnancies resulted in adverse outcomes, including IUD, miscarriages or neonatal death. A recent review on listeriosis during pregnancy also highlighted the significant adverse impact of *Listeria* infection on pregnancy outcomes, including stillbirth, intrauterine fetal demise, spontaneous abortion, neonatal listeriosis, and preterm birth [23]. This underscores the importance of educating pregnant women about preventive measures, raising awareness among healthcare professionals, including obstetricians, regarding early diagnosis and management of listeriosis during pregnancy to improve maternal and fetal outcomes.

Neurolisteriosis was also common in our study, with most patients experiencing meningitis or meningoencephalitis, and only one patient presenting with a brain abscess. The MONALISA group also reported that, among patients with neurolisteriosis, meningoencephalitis was the predominant presentation (84%), and brainstem involvement was observed in only 17%. In the subgroup analysis, no statistically significant difference was found between patients with and without CNS involvement regarding demographic characteristics, comorbidities, clinical presentations, and outcomes. CNS involvement appears to be less common among pregnant females. The incidence of neurolisteriosis reported in the literature is 30% but the occurrence of cerebral abscess secondary to monocytogenes is only 3%.[24].Our patient with listeria brain abscess was an elderly female with a history of diabetes mellitus, hypertension, non-B non-C chronic liver disease and showed complete recovery after 6 weeks of antibiotics . There is limited evidence available, primarily derived from case reports, about the management of listeria brain abscess. Prolonged antibiotic therapy (5-6 weeks) appears reasonable, with variable results of adjunctive surgical drainage. [25-27]

A recent meta-analysis reported a listeriosis-related mortality rate of approximately 23%, with most deaths occurring in individuals over 60 years of age and found that gender has no impact on mortality risk.[10]. The risk factors of mortality associated with listeriosis are neurolisteriosis, prior comorbidities like non-haematological malignancies, pulmonary diseases, chronic kidney diseases, cardiovascular diseases, malignancies, immunosuppression and multi-organ failure. [5, 10, 28].We also found high mortality among the elderly population, with no significant difference between males and females. All except one had existing comorbid illness or immunosuppressive state, but neurolisteriosis was not identified as a risk factor of mortality in the current study. This might be due to small sample size and timely management of patients presenting with neurological involvement, i.e. prompt diagnostic evaluation and early initiation of antibiotics in this high-risk group.

Our study has several limitations. It is a single-centre, retrospective study with a small sample size, and large-scale multicenter studies are required to validate our findings. It was a hospital-based study; many patients with less severe infections may have been managed in primary health centres and might not have presented to tertiary care hospitals, potentially leading to underrepresentation of milder cases. Additionally, retrospective design may have resulted in incomplete documentation with missing information on predisposing factors such as dietary history and environmental exposures.

## CONCLUSION

Listeriosis significantly affects not only the elderly but also middle-aged individuals, particularly those with underlying risk factors and during pregnancy. Environmental hygiene, early diagnosis, and prompt treatment are crucial to reducing adverse outcomes, especially in pregnancy-related cases. Raising public awareness, educating healthcare workers for early diagnosis and management, and implementing preventive measures at the community level can greatly improve listeriosis outcomes.

## Data Availability

All data produced in the present work are contained in the manuscript

## Acknowledgements

**None**

## Funding

This research did not receive any specific grant from funding agencies in the public, commercial, or not-for-profit sectors.

## A competing interests

None

## Conflict of interest

None

## Authorship

All authors have made substantial contributions to all of the following:

1. **ZN, IK, IA, TM, KH:** The conception and design of the study, or acquisition of data, or analysis and interpretation of data
2. **ZN, IK, IA, TM**,**KH**: drafting the article or revising it critically for important intellectual content :
3. **ZN, IK, IA**,**TM, KH**: final approval of the version to be submitted

## Notes

### Competing Interest Statement

The authors have declared no competing interest.

### Funding Statement

This study did not receive any funding

### Author Declarations

Ethics Review committee of Aga Khan university hospital gave ethical approval for this work

